# Recent SARS-CoV-2 infection abrogates antibody and B-cell responses to booster vaccination

**DOI:** 10.1101/2022.08.30.22279344

**Authors:** Clarisa M. Buckner, Lela Kardava, Omar El Merhebi, Sandeep R. Narpala, Leonid Serebryannyy, Bob C. Lin, Wei Wang, Xiaozhen Zhang, Felipe Lopes de Assis, Sophie E.M. Kelly, I-Ting Teng, Genevieve E. McCormack, Lauren H. Praiss, Catherine A. Seamon, M. Ali Rai, Heather Kalish, Peter D. Kwong, Michael A. Proschan, Adrian B. McDermott, Anthony S. Fauci, Tae-Wook Chun, Susan Moir

**Affiliations:** Laboratory of Immunoregulation, National Institute of Allergy and Infectious Diseases, National Institutes of Health, Bethesda 20892 MD, USA; Vaccine Research Center, National Institute of Allergy and Infectious Diseases, National Institutes of Health, Bethesda, MD 20892, USA; Bioengineering and Physical Sciences Shared Resource, National Institute of Biomedical Imaging and Bioengineering, National Institutes of Health, Bethesda, MD 20892, USA; Critical Care Medicine Department, Clinical Center, National Institutes of Health, Bethesda 20892 MD, USA; Biostatistics Research Branch, National Institute of Allergy and Infectious Diseases, National Institutes of Health, Bethesda, MD 20892, USA

**Keywords:** SARS-CoV-2, mRNA vaccines, booster vaccination, memory B cells, antibodies, hybrid immunity, variants, infection

## Abstract

SARS-CoV-2 mRNA booster vaccines provide protection from severe disease, eliciting strong immunity that is further boosted by previous infection. However, it is unclear whether these immune responses are affected by the interval between infection and vaccination. Over a two-month period, we evaluated antibody and B-cell responses to a third dose mRNA vaccine in 66 individuals with different infection histories. Uninfected and post-boost but not previously infected individuals mounted robust ancestral and variant spike-binding and neutralizing antibodies, and memory B cells. Spike-specific B-cell responses from recent infection were elevated at pre-boost but comparatively less so at 60 days post-boost compared to uninfected individuals, and these differences were linked to baseline frequencies of CD27^lo^ B cells. Day 60 to baseline ratio of BCR signaling measured by phosphorylation of Syk was inversely correlated to days between infection and vaccination. Thus, B-cell responses to booster vaccines are impeded by recent infection.

## INTRODUCTION

Severe acute respiratory syndrome coronavirus 2 (SARS-CoV-2) mRNA vaccines provide protection against symptomatic infection through the induction of strong humoral and cellular immunity (Laidlaw and Ellebedy, 2022; Sette and Crotty, 2021). The original two-dose BNT162b2 (Pfizer–BioNTech) or mRNA-1273 (Moderna) vaccine elicits antibodies that are highly effective at neutralizing the ancestral virus (Baden et al., 2021; Polack et al., 2020). More recent studies show booster doses increase potency and breadth of the neutralizing antibody response and the induction of strong CD4^+^ T-cell and memory B-cell responses against variants of concern (VOC) (Goel et al., 2022; Zhang et al., 2022). Boosted immunity as a result of infection and vaccination, commonly referred to as hybrid immunity, is also highly protective against VOC (Bhattacharya, 2022; Laidlaw and Ellebedy, 2022). In a study designed to delineate the effects of mRNA vaccination and/or previous infection on symptomatic infection and severity of disease from Omicron subvariants BA.1 and BA.2, hybrid immunity resulting from previous infection and three doses of vaccine provided the best protection (Altarawneh et al., 2022).

Hybrid immunity from prior infection can provide both quantitative and qualitative benefits by imprinting effector CD4^+^ T-cell populations with enhanced antiviral properties and improving potency and breadth of B-cell and antibody responses (Andreano et al., 2021; Rodda et al., 2022). However, some of these benefits may not extend to booster doses (Rodda et al., 2022), and the effects may be modulated by vaccine and/or infection histories. For example, imprinting from booster vaccination has an attenuating effect on response to Omicron infection while responses to other VOC are boosted and response to Omicron is severely dampened by prior infection with ancestral but less affected by infections with other VOC (Reynolds et al., 2022). Added to the increasing complexities associated with effects of infection and re-infection histories are challenges associated with timing of vaccines and how repeated boosting, whether through vaccination or infection, affects the magnitude and durability of protective immunity. Previous findings from primary two-dose mRNA vaccines suggest that an extended interval between doses increases neutralizing antibody and cellular responses (Payne et al., 2021), especially B-cell responses (Nicolas et al., 2022). However, as exposures to SARS-CoV-2 increase, whether through vaccination, infection, or both, it is unclear how timing between exposures modulates these responses. The risk of deleterious effects on the immune system from repeated and frequent stimulation with the same antigen is known from animal models where antibody-mediated feedback and other regulatory mechanisms have been described (Mesin et al., 2020; Zhang et al., 2013). The role of pre-existing antibody levels in regulating and restricting B-cell responses is also reported in a SARS-CoV-2 mRNA vaccinee plasma transfer model (Dangi et al., 2022).

In the present study, we investigate the effects of SARS-CoV-2 infection on antibody and B-cell responses to a third dose of BNT162b2 or mRNA-1273 vaccine in a longitudinal cohort of uninfected, previously infected, and post-boost infected subjects. While we find robust spike-specific antibody and B-cell responses to the booster vaccine in both uninfected and post-boost infected individuals, responses are muted in those who were infected prior to boosting. We present evidence that the interval between prior infection and booster vaccination is a critical determinant of the immune response to the booster vaccine and that B cells of individuals who were recently infected are unresponsive to the booster vaccine. Our findings thus identify timing relative to infection as a key factor in immune boosting.

## RESULTS

### Prior SARS-CoV-2 infection restricts post-boost binding and neutralizing antibodies

From October 2021 through March 2022, a cohort of 66 adults scheduled to receive a third dose (booster) of either BNT162b2 or mRNA-1273 vaccine was recruited to donate blood at baseline (day 0) and days 30 and 60 post-vaccination. At day 60, participants were stratified into three groups based on exposure to SARS-CoV-2: uninfected (N=44) by nucleocapsid antibody serology, prior-infected (N=11) or post-infected (N=11) depending on whether exposure occurred prior to or after booster vaccination, respectively (Figure 1A and Table 1). In the post-infected group, BA.1 Omicron was the PCR-confirmed or suspected infecting variant while in the prior-infected group, all infections occurred before the first reports of Omicron (data not shown). The interval between vaccination and post-boost infection ranged from 12 to 44 days with a median of 29 days, and the interval between prior infection and vaccination ranged from 59 to 601 days with a median of 160 days (Table 1). Demographics and schedule/source of vaccines did not significantly differ between the groups, except age was slightly lower in the post-infected compared to uninfected but not prior-infected group (Table 1 and Figure S1A).

**Figure 1.**
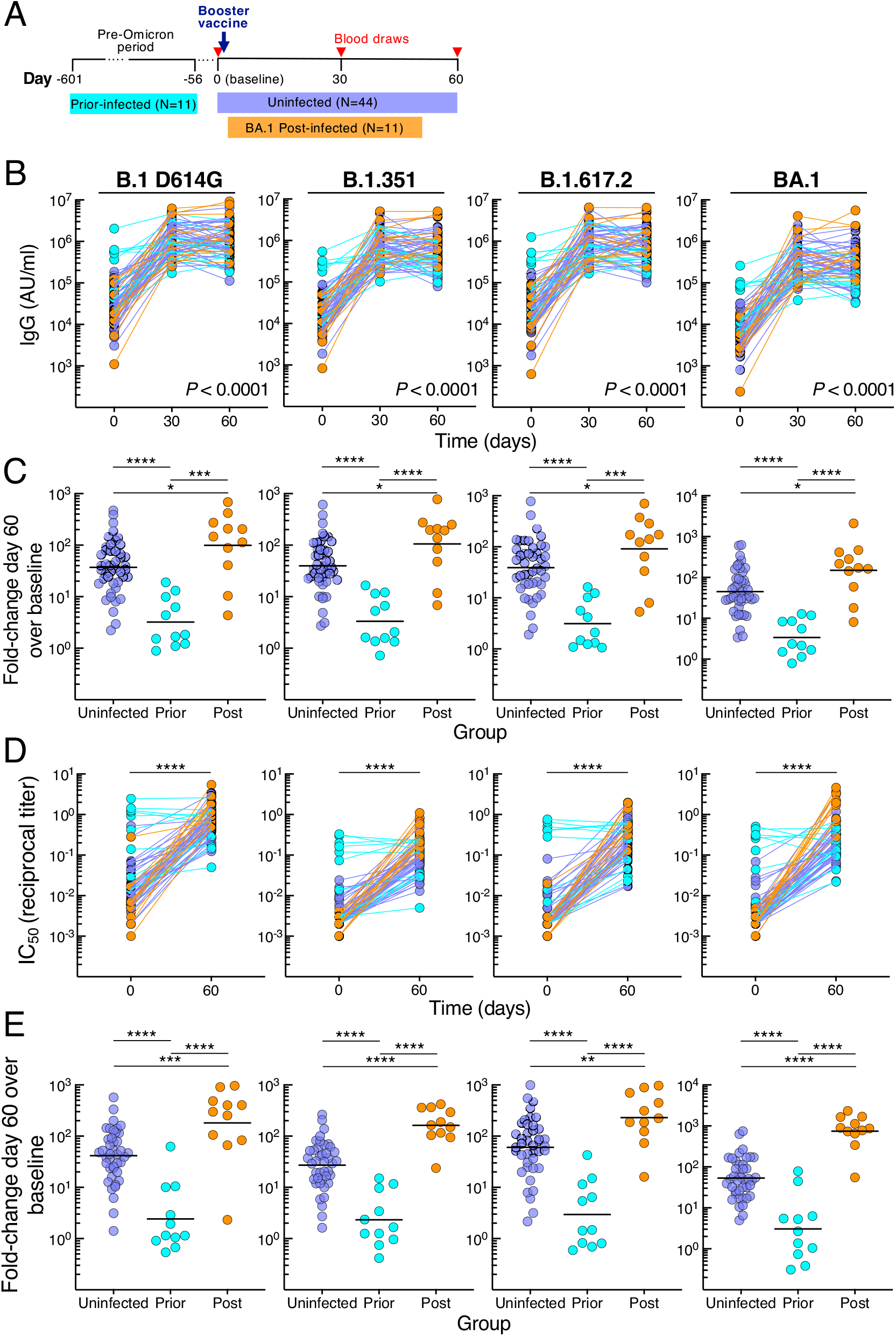
SARS-CoV-2 infection status alters antibody responses to booster vaccination. (A) Graphical depiction of cohort and vaccination/blood draw schedule. (B) IgG spike-binding titers expressed as AU/ml serum. (C) Fold-change day 60 over baseline in serum IgG spike-binding titers. (D) Serum neutralizing titers against SARS-CoV-2 lentiviral pseudovirus expressed as reciprocal IC_50._ N=41 for uninfected group. (E) Fold-change day 60 over baseline in serum neutralizing titers against SARS-CoV-2 lentiviral pseudovirus. Numerical p-values represent mixed-effect model testing of group by time interactions; see Table S2 for additional p-values. Horizontal lines represent geometrical means and coloring scheme matches groups described in Figure 1A.

**Table 1.** Group characteristics.

|  | **Uninfected group** | **Prior-infected group** | **Post-infected group** |
| --- | --- | --- | --- |
| Subjects (number) | 44 | 11 | 11 |
| Age (years) | 54 (21-70)<sup>a</sup> | 45 (31-65) | 38 (23-56) |
| Female gender (number, %) | 28 (64%) | 8 (73%) | 6 (54%) |
| Prior infection to booster vaccine (days) | N/A | 160 (59-601) | N/A |
| Booster vaccine to post infection (days) | N/A | N/A | 29 (12-44) |
| Asymptomatic cases (number, %) | N/A | 2 (18%) | 3 (27%) |
| Moderna; Pfizer; mixed (number, %) | 37 (84%); 5 (11%); 2 (4%) | 8 (73%); 3 (27%) | 8 (73%); 3 (27%) |
| Vaccine dose 1 to booster (days) | 318 (204-393) | 323 (256-393) | 322 (217-353) |
| Day 30 visit from booster vaccine (days) | 30 (26-32) | 29 (28-32) | 30 (28-36) |
| Day 60 visit from booster vaccine (days) | 60 (55-73) | 61 (56-68) | 60 (56-63) |

<sup>a</sup>Median (range)

Serum immunoglobulin G (IgG) antibodies were measured against spike proteins of ancestral B.1 (D614G mutation) and nine variants, including VOC B.1.1.7 (Alpha), B.1.351 (Beta), B.1.617.2 (Delta; AY.4 lineage), and B.1.1.529 (Omicron; BA.1, BA.2 and BA.3 lineages) using a multiplex platform. Antibody titers increased from baseline to day 30 in all three groups, albeit more modestly for the prior-infected group (Figures 1B and S1B-C). By day 60, titers had plateaued in the uninfected group, increased slightly in the post-infected group against B.1 D614G and three variants, yet decreased in the prior-infected group against B.1 D614G and six variants (Figure S1C and Table S1). These temporal differences between groups, evaluated in a mixed-effects model considering time and exposure status, were highly significant for all three groups combined (*P* < 0.0001 for group by time interaction for each of the 10 spike measurements) and between prior-infected and both uninfected and post-infected (*P* < 0.0001 for each of the 10 spike measurements) but not between uninfected and post-infected groups (Table S2 showing all comparisons). At baseline, titers against all 10 spike proteins were higher in the prior-infected than uninfected and post-infected groups, while day 60 titers were lower in the prior-infected than post-infected and for five variants in the uninfected group (Figure S1D and Table S1). Titers were similar between the uninfected and post-infected group at baseline and marginally higher at day 60 in the post-infected group (Figure S1D and Table S1). The combination of high baseline titers with modest increases post-boost in the prior-infected group translated into fold increases between baseline and day 60 that were much lower compared to those of the uninfected and post-infected groups (Figure 1C and Table S1). As expected, the fold increases were modestly higher in the post-infected compared to the uninfected group (Figure 1C and Table S1).

Serum neutralizing antibody activities were measured against B.1 D614G, B.1.351 B.1.617.2 and BA.1 using a pseudovirus-based assay (Gagne et al., 2022). Consistent with the antibody binding titers, neutralization increased from baseline to day 60 in the uninfected and post-infected groups (Figures 1D and S1E), and baseline titers were distinctly elevated in the prior-infected group (Figures 1D and S1F). However, in contrast to antibody binding, neutralizing titers did not increase between baseline and day 60 in the prior-infection group (Figure S1E). Baseline neutralizing titers against B.1.351 and BA.1 were modestly lower in the post-infection compared to uninfected group but were higher at day 60 (Figure S1F). These differences translated into higher fold increases between baseline and day 60 in the post-infected group compared to the two other groups and higher in the uninfected than prior-infected group (Figure 1E), consistent with the antibody binding data. Overall, the kinetics and magnitude of antibody binding and neutralizing titers differed between the groups, although most strikingly in the prior-infected group where antibody responses to the booster vaccine were muted.

### Prior SARS-CoV-2 infection restricts post-boost SARS-CoV-2 B-cell responses

We used spectral flow cytometry to evaluate B-cell responses to the booster vaccine with a 21-marker panel that included six tetramers for evaluating responses against the spike protein subunit 1 (S1) of B.1, its receptor binding domain (RBD) and N-terminal domain (NTD), as well as identifying B cells that bound RBD and NTD of both B.1 and BA.1 (Figures 2A and S2A). Frequencies of B.1 RBD, NTD, as well as dual B.1/BA.1 RBD and NTD-binding B cells increased significantly from baseline to day 60 in the uninfected and post-infected groups, peaking at day 30 in the uninfected group while continuing to increase (RBD) in the post-infected group (Figures 2B and S2B). In contrast, frequencies of spike-binding B cells in the prior-infected group did not significantly increase between baseline and day 30 and declined between days 30 and 60 against B.1 RBD and NTD and B.1/BA.1 NTD (Figures 2B and S2B). These temporal differences between groups, evaluated with the mixed-effects model described above, were highly significant for all three groups combined (*P* < 0.0001 for all five measurements), as well as between group pairs (Table S3 showing all comparisons). As with antibody titers, baseline frequencies of spike-binding B cells were higher in the prior-infected than uninfected and post-infected groups for B.1 RBD and NTD and B.1/BA.1 RBD (Figure S2C). At day 60, spike-binding frequencies were similar between the prior-infected and uninfected group yet were lower for B.1 RBD and B.1/BA.1 RBD in both groups when compared to the post-infected group (Figure S2C). These temporal differences translated into higher fold increases between baseline and day 60 in the post-infected group compared to the two other groups and higher in the uninfected than prior-infected group for both RBD and NTD and dual counterparts (Figure 2C).

**Figure 2.**
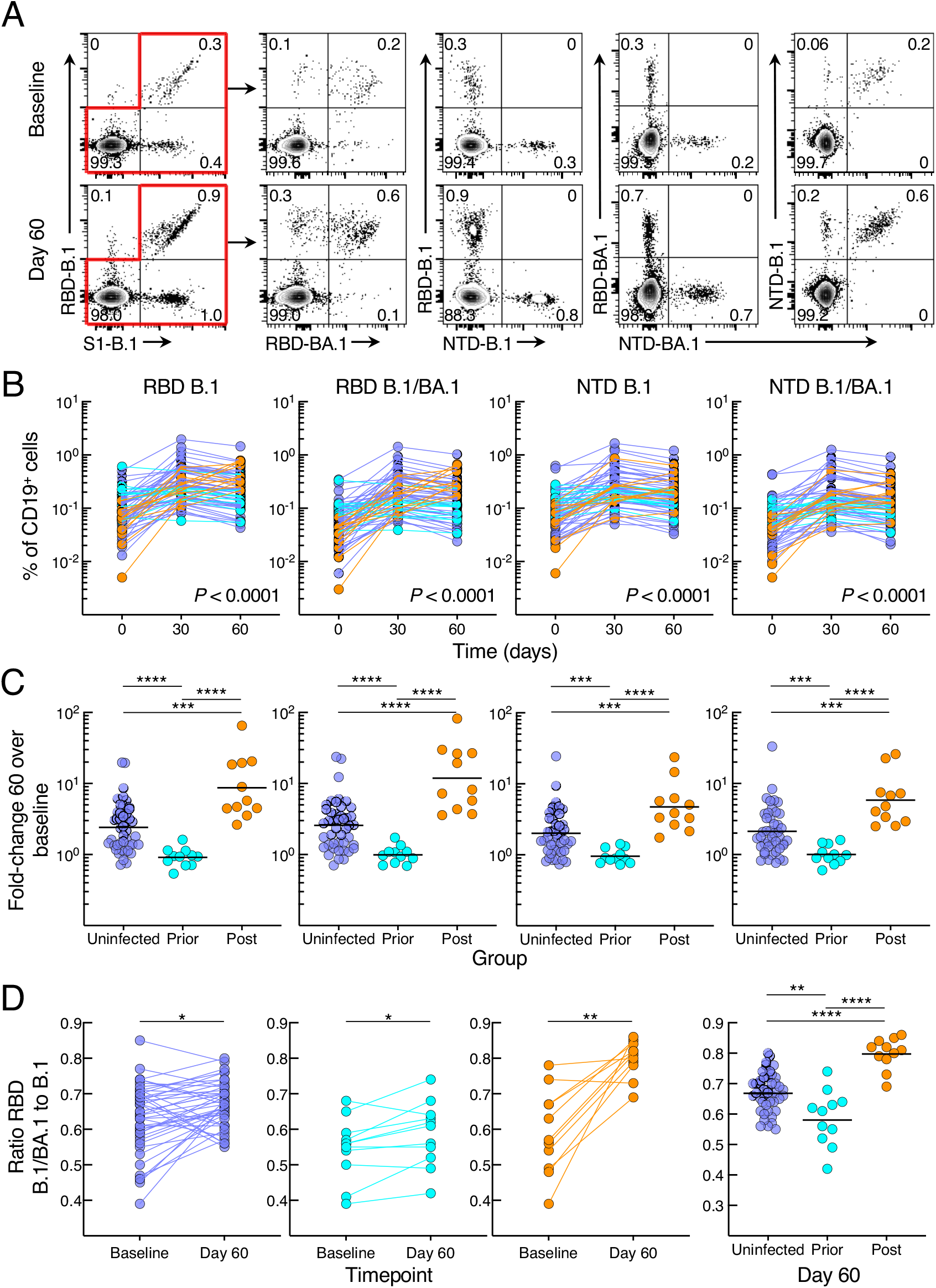
SARS-CoV-2 infection status alters B-cell responses to booster vaccination. (A) Representative binding of B.1 and BA.1 RBD and NTD tetramers to non-naïve B cells at baseline and day 60 post booster vaccination. (B) Frequencies of B.1 and BA.1 RBD and NTD tetramer-binding B cells. (C) Fold-change day 60 over baseline of B.1 and BA.1 RBD and NTD tetramer-binding B-cell frequencies. (D) Ratio of dual B.1/BA.1 to single B.1 RBD binding to B cells. Binding frequencies represent percentage of total CD19^+^ B cells; see also Figure S2A for gating strategy. Numerical p-values represent mixed-effect model testing; see Table S3 for additional p-values. Horizontal lines represent geometrical means and coloring scheme matches groups described in Figure 1A.

We then considered whether post-boost BA.1 infection and/or the booster vaccine altered the ratio of dual B.1/BA.1 to single B.1 RBD or NTD binding. For RBD, the ratio of dual B.1/BA.1 to single B.1 binding increased from baseline to day 60 in all three groups; however, the ratio was highest in the post-infected group and lowest in the prior-infected group (Figure 2D). For NTD, the ratios increased from baseline to day 60 in the uninfected and post-infected but not prior-infected group and at day 60, and the ratios at day 60 were similarly higher in the uninfected and post-infected than in the prior-infected group (Figure S2D). Overall, the B-cell response to the booster vaccine was poor in magnitude and breadth in the prior-infected group while robust in the uninfected and post-infected groups, with the latter having an additional boost from the breakthrough infection.

### Prior SARS-CoV-2 infection restricts post-boost memory B-cell derived antibodies

To gain further insight into B-cell VOC responses after booster vaccination and the effect of post-boost BA.1 infection, we performed *in vitro* culturing of peripheral blood mononuclear cells (PBMCs) at baseline and day 60 using polyclonal stimuli known to favor memory B-cell differentiation and antibody secretion (Goel et al., 2021). We measured culture-derived antibodies with the same multiplex platform used for serum IgG titers, although relative activity levels were typically 2-3 logs lower in cultures than in serum and reported per total secreted IgG (Figure 3A). Consistent with the spike-binding B-cell kinetics, spike-specific antibodies secreted from cultured PBMCs increased between baseline and day 60 in the uninfected and post-infected but not prior-infected group (Figures 3A and S3A-B and Table S4). These temporal differences, evaluated with the mixed-effects model described above, were highly significant for all three groups combined (*P* < 0.0001 for each of the 10 spike measurements), between prior-infected and both uninfected and post-infected groups (*P* < 0.0001 for each of the 10 spike measurements), as well as between the uninfected and post-infected group, albeit somewhat less pronounced (Table S5 showing all comparisons). Baseline secreted antibodies were modestly higher in the prior-infected group compared to uninfected and/or post-infected group against the five non-Omicron spike proteins whereas there were no baseline differences against the five Omicron variants tested (Figure S3C and Table S4), the lack of measurable differences possibly due to low baseline Omicron-specific antibodies. However, by day 60, secreted antibody titers were lower in the prior-infected compared to both uninfected and post-infected group, and lower in the uninfected compared to the post-infected group (Figure S3C and Table S4). As with serum antibodies and spike-binding B cells, these temporal differences translated into higher fold increases of secreted antibodies between baseline and day 60 in the post-infected group compared to the two other groups and higher in the uninfected than prior-infected group against all 10 spike proteins (Figure 3B and Table S4). Overall, spike-specific antibodies secreted from memory B cells did not increase in the prior-infected group after the vaccine boost and while levels compared to both uninfected and post-infected groups were similar or modestly higher at baseline, they were sharply lower by day 60.

**Figure 3.**
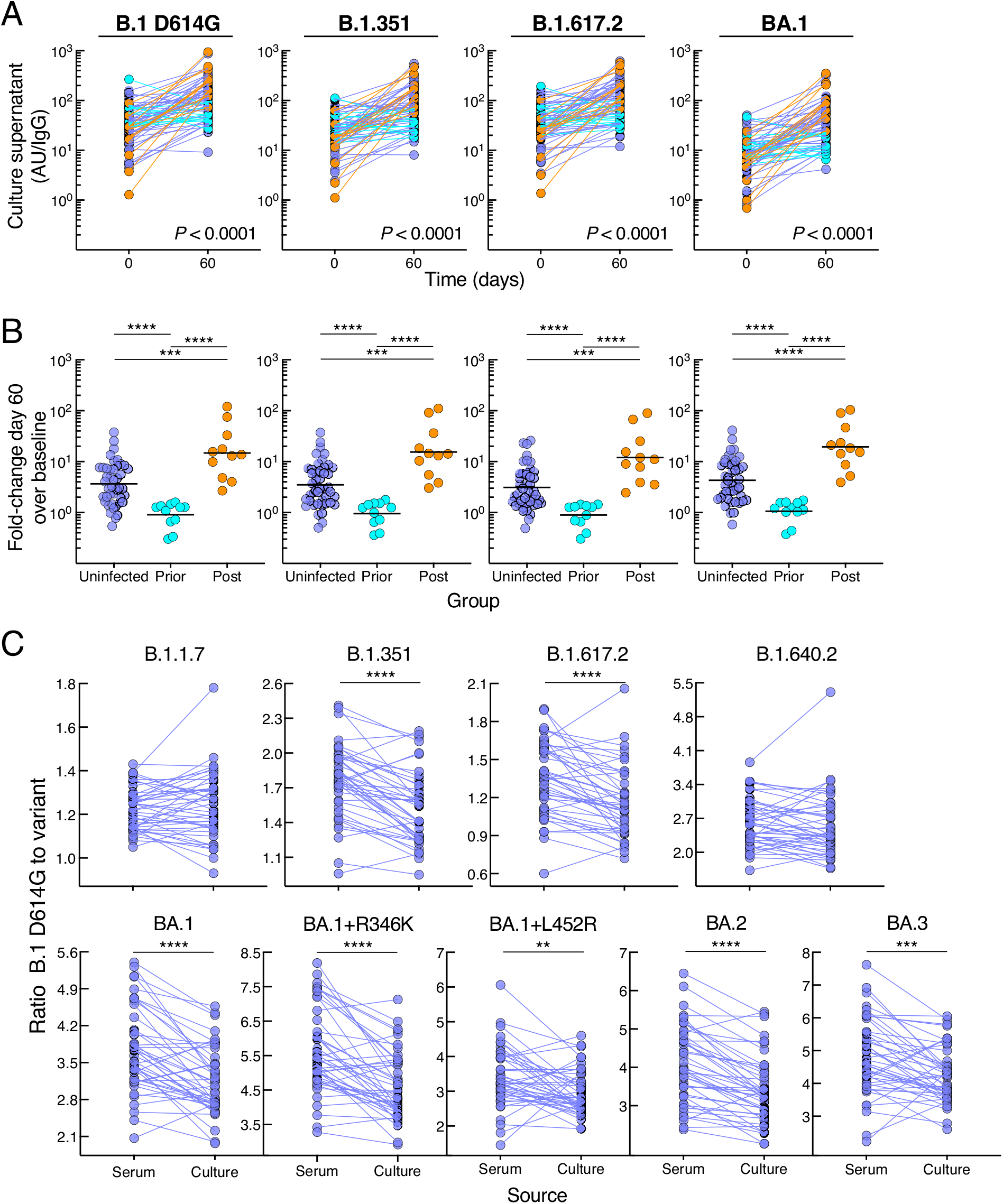
SARS-CoV-2 infection status alters secreted antibody responses to booster vaccination. (A) IgG spike-binding titers in culture supernatant expressed as AU/IgG. (B) Fold-change day 60 over baseline of IgG spike-binding titers in culture supernatant. (C) Ratio of B.1 D614G to variant IgG spike-binding titers in serum and culture supernatant. Culture supernatant titers represent spike-binding IgG per total IgG. Numerical p-values represent mixed-effect model testing; see Table S5 for additional p-values. Horizontal lines represent geometrical means and coloring scheme matches groups described in Figure 1A.

### Enhanced breadth of antibodies secreted from memory B cells

The robust secreted antibody response at day 60 in the large uninfected group provided an opportunity to consider whether antibodies derived from memory B cells had greater breadth compared to serum antibodies, as suggested in previous studies (Muecksch et al., 2022; Purtha et al., 2011). Accordingly, we compared ratios of B.1 D614G to variant antibody titers between the two sources. For seven of the nine spike variants tested, the fold difference was significantly reduced for the culture-secreted antibodies compared to those in serum while differences were not significant for two variants (Figure 3C), confirming the superior breadth in memory B-cell compartment.

### Prior SARS-CoV-2 infection alters B-cell responses and phenotypes

The fold changes in memory B-cell responses to booster vaccination were significantly lower in the prior-infected group compared to the two other groups, whether measured by direct binding (Figure 2) or secreted antibodies (Figure 3). However, at day 60, titers of secreted spike-binding antibodies were significantly lower in the prior-infected compared to the uninfected group (Figure S3C), while frequencies of spike-binding B cells were similar (Figure S2C). This contrast suggested that prior infection may have led to a reduced B-cell responsiveness to stimulation after vaccination. We also observed a dichotomous pattern of antibody titers and B-cell responses within the prior-infected group (clearly discernable in Figures 1B and S1B-C) that may have been driven by the wide range of intervals between infection and vaccination (59-601 days: Table 1). This led us to consider whether the interval between infection and vaccination may have contributed to the differences in antibody and B-cell responses observed within this group. Notably, in individuals whose infection occurred more than 180 days prior to receiving their booster vaccine, secreted spike-binding antibodies increased between baseline and day 60 (Figure S4A), as observed for most individuals in the uninfected and all in the post-infected group (Figure S3B). In contrast, all decreases in secreted antibodies between baseline and day 60 were among the six individuals whose breakthrough infection occurred less than 180 days from vaccination (Figure S4A).

To determine whether the reduced antibody secretion in the prior-infected group reflected lower B-cell proliferation and differentiation *in vitro*, we cultured CFSE-labeled baseline and day 60 PBMCs of the six individuals whose prior infection was < 180 days from vaccination and of six uninfected individuals matched for demographics and baseline B-cell spike-binding frequencies. After four days in culture under the same conditions used to induce antibody secretion, the cells were stained with RBD and S1 tetramers and CFSE dilution was measured among spike-positive (RBD^+^S1^+^) and spike-negative (RBD^-^S1^-^) IgG-expressing B cells (Figures 4A and S4B). The division index, a measure of cell division among both proliferating and non-proliferating cells, did not significantly differ at baseline and day 60 within and between the two groups for both RBD^+^S1^+^ and RBD^-^S1^-^ B cells, although there was a clear downward trend among RBD^+^S1^+^ B cells of five of the six individuals in the prior-infected group (Figure 4B). This trend translated into a ratio of day 60 to baseline indices for RBD^+^S1^+^ B cells that was significantly lower in the prior-infected compared to the uninfected group, while ratios among RBD^-^S1^-^ B cells remained close to 1.0 for both groups and for RBD^+^S1^+^ B cells of the uninfected group (Figure 4C). Notably, the individual in the prior-infected group with an RBD^+^S1^+^ B-cell ratio above 1.0 in Figure 4C (denoted by gray circles) was one of two, and the higher of the two in the subgroup, whose secreted spike-binding antibodies increased between baseline and day 60 (Figure S4A).

**Figure 4.**
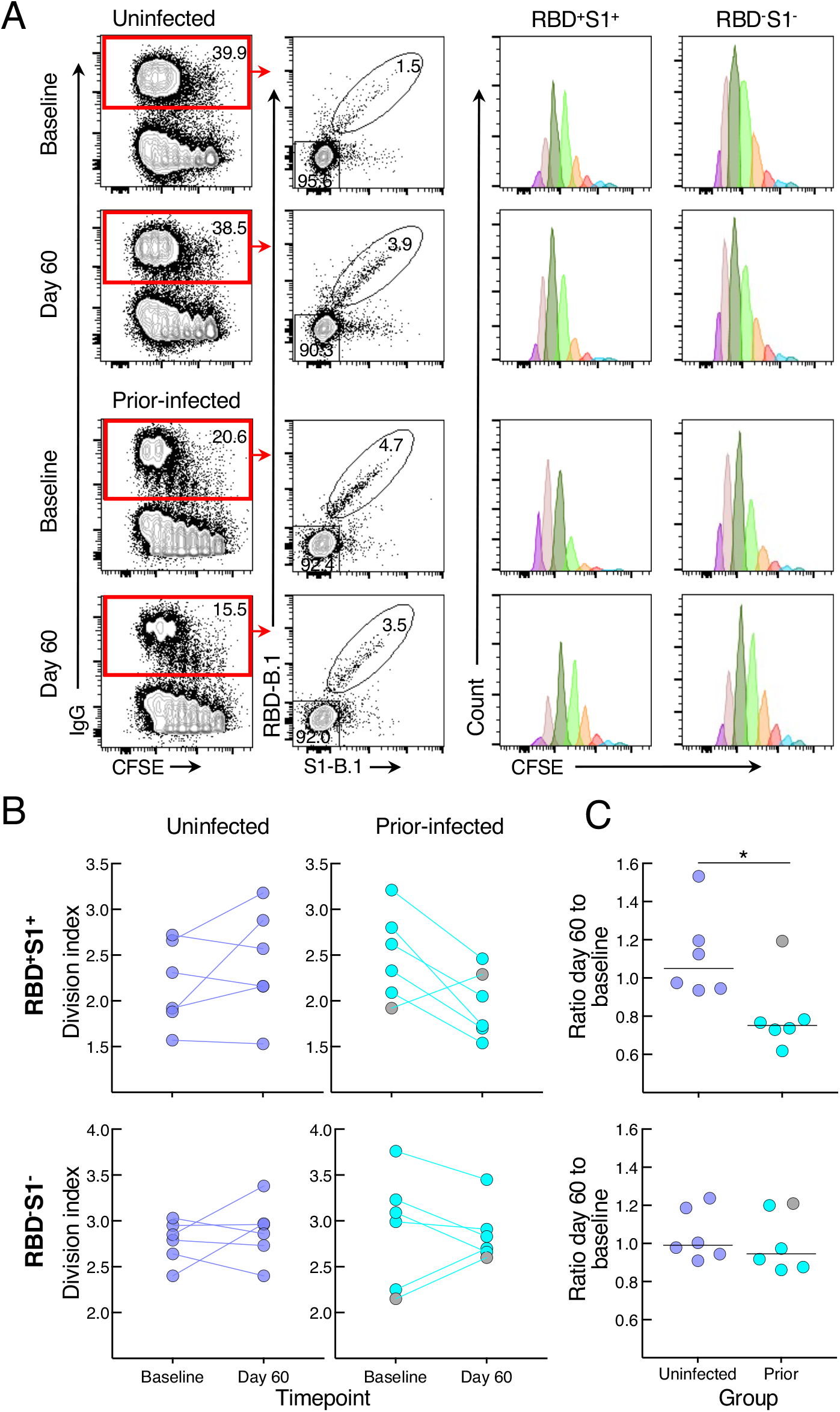
SARS-CoV-2 recent infection decreases proliferative response of spike-specific B cells after booster vaccination. (A) Representative baseline and day 60 CFSE-dilution plots of RBD^+^S1^+^ and RBD^-^S1^-^ IgG-expressing B cells among stimulated PBMCs. Numbers represent percentages. (B) Division indices of RBD^+^S1^+^ and RBD^-^S1^-^ IgG-expressing B cells. (C) Ratio of day 60 to baseline division indices of RBD^+^S1^+^ and RBD^-^S1^-^ IgG-expressing B cells. CFSE-labeled PBMCs were stimulated for four days then stained with anti-IgG, B.1 S1 and RBD tetramers and other markers shown with gating strategy in Figure S4B. Division indices were measured for RBD^+^S1^+^ and RBD^-^S1^-^ IgG-gated B cells. Horizontal lines represent medians and coloring scheme matches groups described in Figure 1A. Gray circles refer to subject shown in Figure S4A.

We then considered whether the relative reduction in responsiveness of cultured spike-specific B cells in the prior infection sub-group was intrinsic to the B cells. While there are many assays to assess B-cell responsiveness, including intracellular calcium flux and signaling responses following B-cell receptor (BCR) stimulation, only the latter can adequately capture rare antigen-specific events within the confines of cells available in clinical trials and the narrow timeframe needed to detect bound antigen. Accordingly, we stimulated the B cells with anti-BCR and measured phosphorylation of molecules known to undergo rapid and strong induction following antigen binding and BCR aggregation (Packard and Cambier, 2013). Preliminary assays identified the tyrosine kinase Syk and phospholipase Cγ2 (PLCγ2), which lies downstream of Syk, as being rapidly phosphorylated and clearly discernable among the RBD-binding B cells following anti-BCR treatment (Figure 5A). In the uninfected group, BCR signaling led to similar levels of phosphorylated (p-) Syk, p-PLCγ2 and combination of p-Syk/p-PLCγ2 RBD^+^ B cells at baseline and day 60, while in the prior-infected group levels for p-Syk and p-Syk/p-PLCγ2 decreased between baseline and day 60 and baseline levels were higher than in the uninfected group for all three measurements (Figure 5B). Among RBD^-^ B cells, there was a downward trend in the prior-infected group between baseline and day 60 that was significant for p-PLCγ2, although there were no differences between the groups (Figure S5A). These temporal dynamics translated into ratios of day 60 to baseline p-Syk and p-Syk/p-PLCγ2 that were close to 1.0 among RBD^-^ and RBD^+^ B cells of the uninfected group while they were below 1.0 among RBD^+^, and to a lesser extent, RBD^-^ B cells of the prior-infected group and significantly lower than those of the uninfected group (Figure 5C). For p-PLCγ2, ratios were close to 1.0 and not significantly different between the groups (Figure S5B). Furthermore, within the prior-infection group, the interval between infection and vaccination was strongly correlated with the p-Syk ratio in that the shorter the interval the lower the induction of p-Syk at day 60 relative to baseline (Figure 5D).

**Figure 5.**
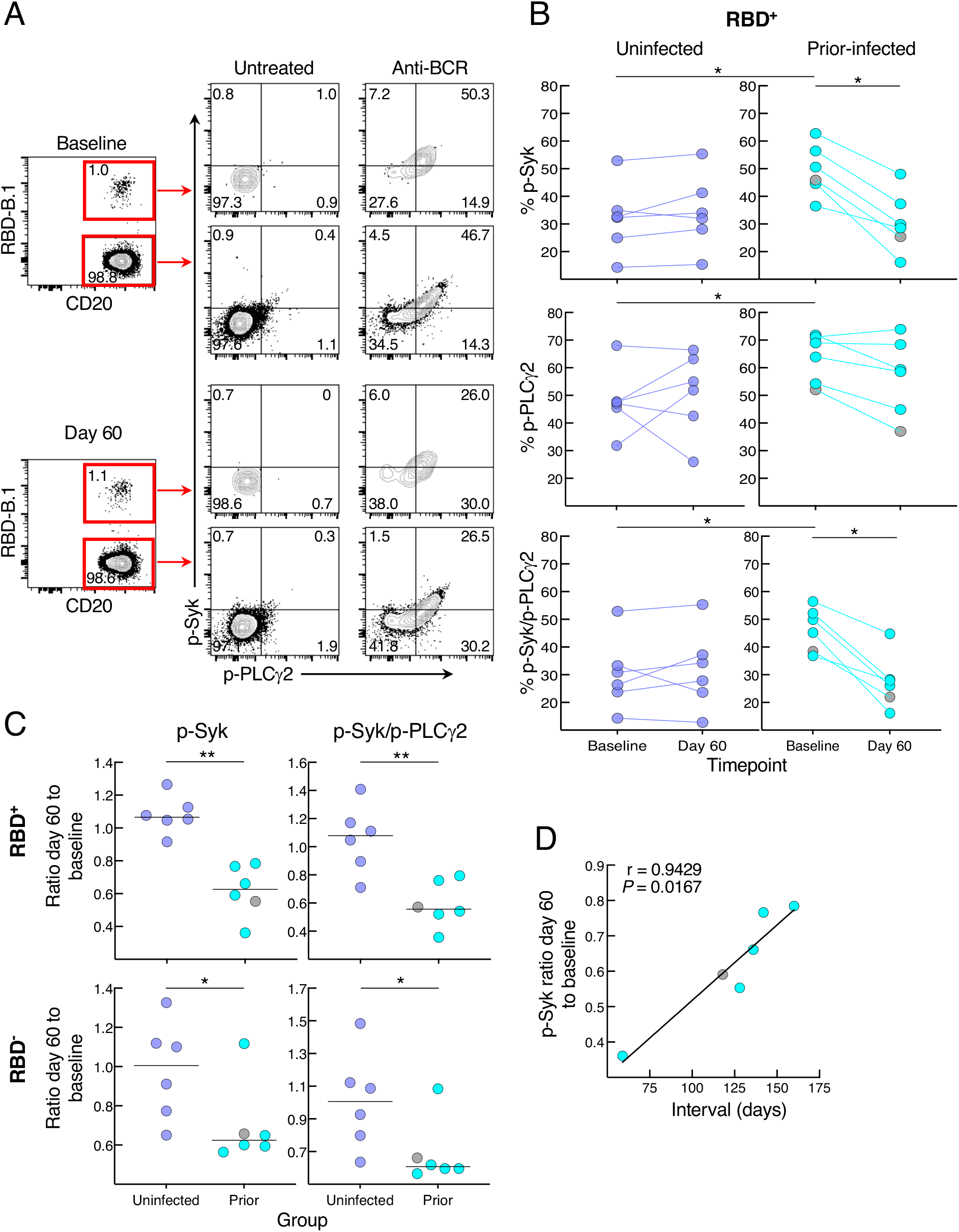
SARS-CoV-2 recent infection decreases BCR signaling response of spike-specific B cells after booster vaccination. (A) Representative baseline and day 60 phosphorylation of Syk and PLCγ2 with and without anti-BCR treatment shown for RBD^+^ and RBD^-^ non-naïve B cells of a recent prior-infected subject. Numbers represent percentages. (B) Anti-BCR induced phosphorylation of Syk and PLCγ2 among RBD^+^ B cells, showing untreated subtracted frequencies. (C) Ratio of day 60 to baseline of anti-BCR induced phosphorylation of Syk and Syk/PLCγ2 among RBD^+^ and RBD^-^ B cells. (C) Correlation of p-Syk ratio day 60 to baseline and time interval between infection and vaccination. PBMCs were stained with B.1 RBD tetramer, IgD and CD27 for gating on non-naïve B cells and other markers (see Figure S2B) and incubated at 37°C for two minutes with or without anti-BCR, then stained for phosphorylated markers shown. Horizontal lines represent medians and coloring scheme matches groups described in Figure 1A. Gray circles refer to subject shown Figure S4A.

Elevated basal p-Syk and reduced induction following BCR stimulation are hallmarks of certain chronic conditions such as autoimmunity and persisting pathogens, such as HIV, and are generally associated with distinct B-cell populations, many of which express reduced levels of CD21 (Schrezenmeier et al., 2019). Activation of B cells in response to SARS-CoV-2 infection and vaccination is also accompanied by a reduction in the expression of CD21 on B cells (Kardava et al., 2022; Rodda et al., 2022; Woodruff et al., 2020). Antigen-experienced B cells can also express variable levels of CD27 (Baumgarth, 2021). Both markers were differentially modulated by prior infection on RBD^+^ B cells (Figure 6A). At baseline, the two groups had similarly low frequencies of CD21^lo^ B cells while the uninfected group had higher frequencies of CD21^+^CD27^lo^ B cells (Figure 6B). Between baseline and day 60, frequencies of CD21^lo^ B cells increased in both groups but the increase was greater in the uninfected group (Figure 6B). In contrast, frequencies of CD21^+^CD27^lo^ B cells decreased between baseline and day 60 in the uninfected but not prior-infected group and frequencies were similar at day 60 in the two groups. Finally, we considered whether CD27^lo^ among CD21^+^ B cells were less responsive to BCR stimulation than their CD27^+^ counterparts, and possibly explaining the lower baseline p-Syk and p-PLCγ2 induction in the uninfected compared to the prior-infected group (Figure 5B). At baseline in the uninfected group, frequencies of BCR-induced p-Syk among RBD^+^ B cells were lower for the CD27^lo^ compared to the CD27^+^ fraction (Figure 6C). Collectively, these findings suggest that prior to booster vaccination, spike-specific B cells of recently infected individuals are more poised to respond to stimulation than those of uninfected individuals yet are refractory to further stimulation following vaccination.

**Figure 6.**
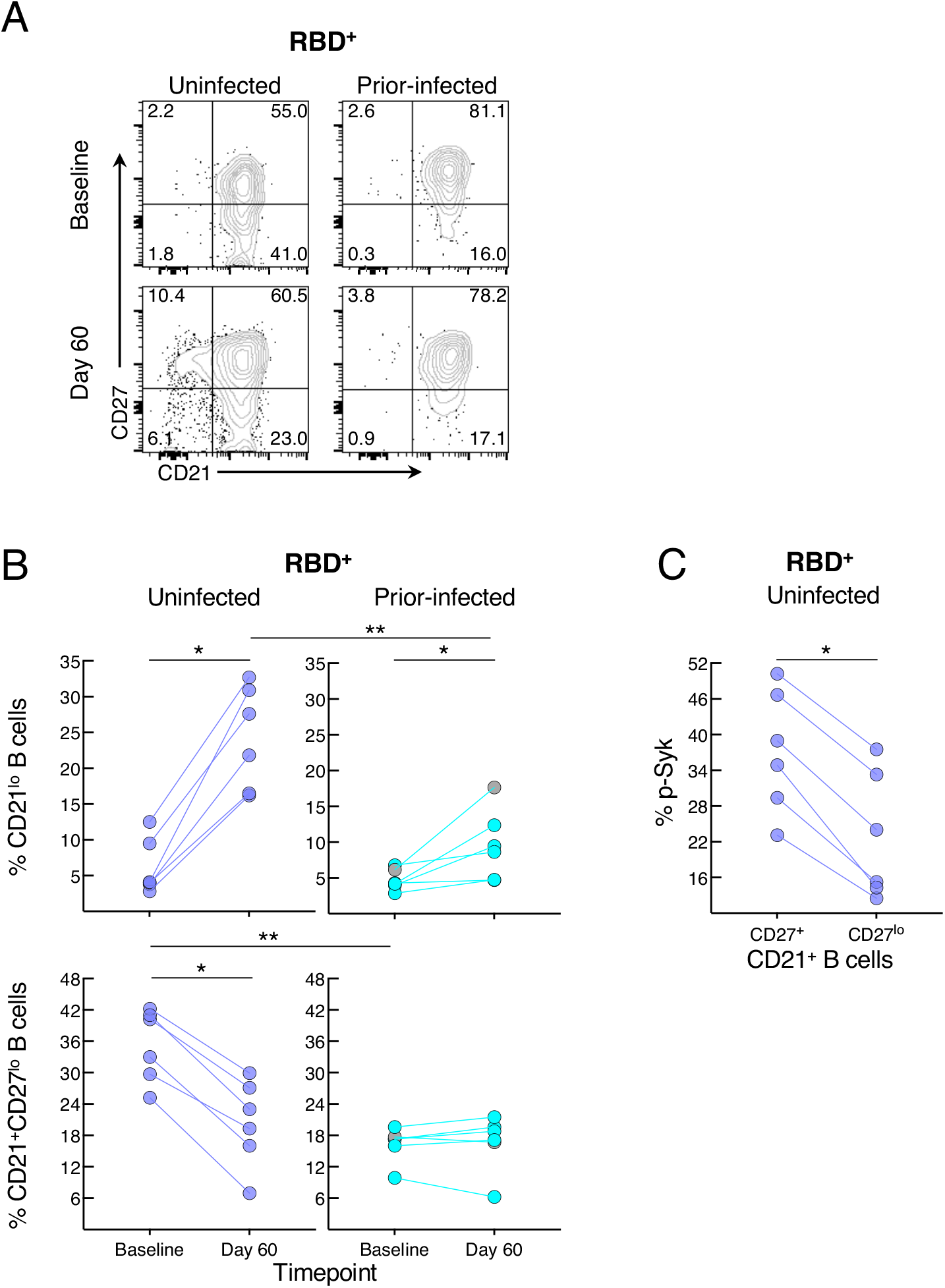
SARS-CoV-2 recent infection alters the phenotype of spike-specific B cells. (A) Representative expression of CD21 and CD27 among RBD^+^ B cells at baseline and day 60 post-boost. Numbers represent percentages. (B) Frequencies of CD21^lo^ and CD21^+^CD27^lo^ RBD^+^ B cells at baseline and day 60 post-boost. (C) Baseline uninfected anti-BCR induced phosphorylation of Syk among CD21^+^CD27^lo^ and CD21^+^CD27^+^ RBD^+^ B cells. Populations shown in (A) and frequencies measured in (B) were from the CD19^+^ gate shown in Figure S2A and those in (C) further gated on CD21^+^ B cells. Horizontal lines represent medians and coloring scheme matches groups described in Figure 1A. Gray circles refer to subject shown in Figure S4A.

## DISCUSSION

Booster vaccines against SARS-CoV-2 play an essential role in protecting against severe COVID-19 (Bhattacharya, 2022). Booster mRNA vaccines induce potent antibody and memory B-cell responses (Goel et al., 2022), although both are modulated by infection histories (Reynolds et al., 2022). In this study, we investigated the effect of SARS-CoV-2 infection on antibody and B-cell responses to mRNA booster vaccines in a longitudinal cohort that included subjects who remained uninfected throughout the study period, as well as subjects who were infected prior to study and subjects who experienced a breakthrough infection during the study period. Collectively, our data identified prior infection, and more specifically, recent prior infection with SARS-CoV-2 to be associated with a diminished response to the booster vaccine when compared to uninfected and post-boost infected subjects. The diminished response in the prior-infected group was most notable for the fact that neutralizing antibodies and B-cell responses to the vaccine boost did not increase between baseline and study endpoint (day 60). Antibody and B-cell responses tended to be higher at baseline in the prior-infected group compared to the two other groups, consistent with the effects of hybrid immunity (Reynolds et al., 2022; Rodda et al., 2022); however, they were similar to or lower than the other two groups at endpoint. Given that vaccine-specific antibody titers and B-cell frequencies were generally similar between the uninfected and prior-infected group, our data do not suggest diminished immunity in prior-infected individuals but rather, that the booster vaccine was ineffective when administered after a recent infection.

We found strong concordance between the antibody and B-cell responses after boosting, both in terms of kinetics, response to variants and differences in responses between the groups. The patterns of fold-change between baseline and day 60 were almost identical from one assay to another, whether serum binding or neutralizing antibody titers or frequencies of spike-specific B cells and their secreted antibodies were measured: the post-infected group experienced the largest fold-change, as might be expected, while the prior-infected group had the lowest fold-change. The temporal dynamics were also similar across the different assays, and these were clearly influenced by infection status, although differences were more pronounced between prior-infected and both uninfected and post-infected groups than between the latter two groups.

Despite the many similarities between the assays, there were notable distinctions. Serum binding antibodies in the prior-infected group did increase after boosting and remained higher at day 60 than baseline, in contrast to all other measurements of antibody and B-cell activity that were unchanged between the two timepoints. It is unclear why the increase was limited to binding antibodies but this may be related to the induction of short-lived plasmablasts that are more likely to produce binding but not more matured neutralizing antibodies (Bhattacharya, 2022). It is noteworthy that binding antibody titers against ancestral and most variants declined between day 30 and 60 in the prior-infected group, but not the uninfected or post-infected groups, consistent with a short-lived plasmablast-mediated rise in the prior-infected group. The temporal dynamics were also different between spike-binding antibodies, which were stable or increased from day 30 to day 60 in groups without prior infection, and spike-binding B cells, which declined during this period in the uninfected group and marginally increased against RBD in the post-infected group. Similar findings have been reported by others and in our previous study (Goel et al., 2022; Kardava et al., 2022), likely reflecting peak B-cell activation that occurs between 2-4 weeks after a second or third dose of vaccine.

Booster vaccination and infection also differentially impacted immune responses to viral variants. As expected, responses to the BA.1 variant in the post-infected group, all of whom were confirmed or suspected to have been infected with BA.1, were strong. However, except for neutralizing antibody titers, the increase in response to the BA.1 spike in the BA.1-infected group was similar in magnitude to responses to ancestral and the other variants spike antigens. The somewhat modest response to BA.1 may be explained by several factors, including vaccinal imprinting (Reynolds et al., 2022), the relatively mild clinical nature of BA.1 (Sigal, 2022), and the proximity of day 60 to the breakthrough infection, a median of 29 days with a range of 12-44 days. We also noted modest yet significantly lower baseline neutralizing antibody titers against BA.1 and B.1.351 in the post-infected compared to the uninfected group, possibly suggesting a higher susceptibility to BA.1 breakthrough infection. Beyond the effect of BA.1 infection on the overall immune response to a range of variants, we noted that antibodies secreted from cultured PBMCs, which most likely originated from memory B cells, had greater breadth compared to serum antibodies, which most likely originated from lymphoid tissue resident plasma cells. These data are consistent with previous animal and more recent COVID-19 vaccine studies (Muecksch et al., 2022; Purtha et al., 2011), highlighting the critical role of the cellular immune response in a pandemic driven by rapidly evolving variants. We also noted that while both uninfected and post-infected groups increased their post-boost ratio of B cells that bound both B.1 and BA.1 RBD and NTD relative to B.1 alone, the increase was restricted to RBD in the prior-infected group, and the ratio at day 60 was significantly lower compared to the other two groups, suggesting a more limited cellular immune response to certain variants.

While all fold-changes between day 60 and baseline revealed a muted post-boost response in the prior-infected group, we were struck by the dichotomous nature of responses within this group and the significantly lower day 60 secreted spike-binding antibody titers despite similar spike-binding B-cell numbers when compared to the uninfected group. We determined that the dichotomous response was largely driven by the interval between infection and booster vaccination, where those who were most recently infected had the highest spike-specific baseline antibody titers and B-cell frequencies, yet the weakest post-boost responses. Given the relatively lower secretion of spike-binding antibodies from *in vitro* stimulated B cells compared to frequencies of spike-binding B cells, we initially considered that recent infection may be causing functional non-responsiveness or post-activation anergy (Schrezenmeier et al., 2019). However, taken collectively, our data suggest a more nuanced effect of recent infection on response to booster vaccination. While spike-specific B cells of recently infected individuals clearly underwent fewer *in vitro* cell divisions at day 60 compared to baseline with a corresponding decrease in BCR signaling, cell division indices and levels of anti-BCR induced phosphorylation were not significantly different at day 60 between the six recently infected and six uninfected individuals. The main differences between these two groups were at baseline, where signaling responses to BCR stimulation were stronger in the recently infected group. We might have expected CD21^lo^ B cells to be responsible for the differences in BCR signaling, given their elevated state of activation/signaling in chronic conditions (Jenks et al., 2018; Kardava et al., 2018), and following vaccination/infection, including in patients with COVID-19 (Castleman et al., 2022; Kardava et al., 2022; Rodda et al., 2022; Woodruff et al., 2020). However, frequencies of CD21^lo^ B cells among spike-specific B cells were found to be similarly low in both groups at baseline and increased at day 60 post-boost in uninfected but not recently infected subjects, consistent with the muted response of the latter group to vaccination. Instead, we found a higher frequency of CD27^lo^ among spike-specific CD21^+^ B cells in the uninfected compared to recently infected group and a corresponding lower BCR signaling response among CD27^lo^ compared to CD27^+^ B cells. Resting CD21^+^CD27^lo^ B cells have been shown to be more durable than other memory B-cell populations following influenza vaccination (Andrews et al., 2019), and were found to increase over time after two doses of the BNT162b2 mRNA vaccine (Kotaki et al., 2022). We speculate that despite being in a resting state, CD27^lo^ B cells are better poised than other memory B cells to respond robustly upon re-encounter with antigen. For the poor responders among the prior-infected group, their infection induced a robust response, as evidenced by the high baseline titers and responding B cells; however, the interval between infection and vaccination was likely insufficient for their B cells to return to an optimal resting state and respond efficiently to the booster.

In summary, we have shown that antibody and B-cell responses to SARS-CoV-2 booster vaccination are impacted by infection status, where prior SARS-CoV-2 infection is associated with a muted response, the extent of which is dictated by the interval between infection and vaccination. When the interval is too short, the response induced by the recent infection appears to prevent B cells from responding to the subsequent booster vaccine. As a growing number of people are infected and re-infected with SARS-CoV-2, these findings may help provide guidance for future recommendations on how to establish booster vaccine schedules that account for infection histories.

### Limitations of the study

There are several limitations to our study, most notably the small number of subjects in the prior-infected and post-infected groups and the limited number of individuals in the former group whose booster vaccine was administered within 180 days of infection. These are inevitable limitations of human studies yet future studies will be needed to confirm and extend our findings, to determine whether additional ancestral and/or variant boosters will lead to similar outcomes in the absence or presence of breakthrough infections. Our study did not consider other likely contributors to outcomes, including the effect of T cells, both *in vivo* and *in vitro* where they, along with other lineages and corresponding soluble factors likely modulated B-cell division, differentiation, and antibody secretion.

## METHODS

### Human study subject recruitment, visits, and clinical data

Sixty-six adults who were scheduled to receive their third dose of the Pfizer (BNT162b2) or Moderna (mRNA-1273) mRNA vaccine were enrolled in an NIH clinical protocol designed for investigating longitudinal vaccine responses to emerging pathogens (ClinicalTrials.gov identifier NCT05078905). The protocol was approved by the NIH Institutional Review Board and written informed consent was obtained from all study subjects. Blood samples were collected at baseline (day 0) prior to (either on the day of or the day before) vaccination, and on day 30 and day 60 post-vaccination. Eleven subjects had COVID-19 prior to and another eleven after receiving their booster vaccine. A summary of demographics as well as infection and vaccine information is provided in Table 1. Forty-four subjects remained uninfected throughout the study period, verified by serum nucleocapsid antibody testing by the Bio-Rad Platelia and a second in-house assay described below.

### Blood sample collection, processing, and storage

PBMCs were isolated by Ficoll density gradient centrifugation from whole blood collected in ethylenediaminetetraacetic acid Vacutainer tubes. Cells were frozen at -80°C and stored in liquid nitrogen. Serum was isolated by centrifugation of clotted whole blood collected in serum separation transport Vacutainer tubes and stored at -80°C.

### *In vitro* differentiation of memory B cells into antibody-secreting cells

PBMCs were thawed, seeded at 1×10^6^ per ml in complete media (RPMI containing 25mM Hepes and L-glutamine + 10% FCS +1% Pen/Strep) and stimulated with 1000 U/ml recombinant human IL-2 (StemCell Technologies) and 2.5 γg/ml R848 (Invivogen) for 7 days at 37°C. On day 4, half the media was removed and replaced with fresh media containing 1X IL-2/R848. Day 7 culture supernatants were collected, spun down to remove cellular debris, assayed for total IgG concentration, and stored at -80°C. Total IgG was measured by cytometric bead array (CBA) assay (BD Biosciences) using a FACS Lyric instrument (BD Biosciences), according to the manufacturer’s instructions. Data were analyzed using BD FACSuite software v1.2.1.5657 (BD Biosciences).

### Serum and culture supernatant antibody titers

Serum samples were heat-inactivated at 56°C for 60 minutes. A 10-plex IgG spike-binding assay was performed on serum and culture supernatant samples with an electrochemiluminescence immunoassay analyzer (ECLIA) developed by Meso Scale Discovery (MSD), as previously described (Kardava et al., 2022), with the following modification. The assays were performed using the MSD V-PLEX SARS-CoV-2 panel 25 kit. The in-house serum nucleocapsid antibody testing was performed using an enzyme-linked immunosorbent assay as previously described (Kalish et al., 2021), with the following adjustments. Nucleocapsid proteins were synthesized as described (Esposito et al., 2020; Mehalko et al., 2021), and longitudinal quality control and assay stability was ensured by the inclusion of recombinant human antibodies on each plate (anti-SARS-CoV-2 RBD IgG, IgM and IgA from GenScript; and, anti-nucleocapsid IgG from ThermoFisher).

### Lentiviral pseudovirus neutralization

Heat-inactivated serum samples were assayed for SARS-CoV-2 neutralization with a lentiviral pseudovirus-based assay, as previously described (Gagne et al., 2022), and reported as reciprocal 50% inhibitory concentration (IC_50_) based on serum IgG µg/ml.

### Biotinylated NTD proteins

The NTD subunit of the spike protein of WA-1 strain (referred to as B.1 in this study) and BA.1 variant were cloned, expressed and biotinylated as previously described (Teng et al., 2022).

### Spike-binding B cells

A 21-color panel was developed to phenotype B-cell populations and identify spike-binding B cells among PBMCs by spectral flow cytometry (Table S6). The biotinylated spike proteins were tetramerized with fluorescently labeled streptavidin (SA) at a molar ratio of 4:1 as described previously (Kardava et al., 2022) with following modifications: S1-B.1 was conjugated with SA-R-Phycoerythrin (PE), RBD-B.1 with SA-BV421, RBD-BA.1 with SA-Allophycocyanin (APC), NTD-B.1 with SA-Alexa Fluor 488, NTD-BA.1 with SA-BUV615 and S-2P-B.1 with SA-PE-Cy5.5. Cryopreserved PBMCs were thawed, and 3 × 10^6^ cells were stained with Zombie NIR Fixable Viability Dye at room temperature for 10 minutes, followed by staining with a cocktail containing 14 monoclonal antibodies (mAbs) and 6 fluorochrome-conjugated spike proteins in staining buffer (2% FBS/PBS) supplemented with Brilliant Stain Buffer Plus (BD Biosciences) at 4°C for 30 minutes. The stained cells were fixed (Lysing Solution, BD Biosciences) and acquired on an Aurora spectral cytometer using SpectroFlo Software v3.0.1 (Cytek Biosciences) and analyzed using FlowJo v10 (BD Biosciences).

### Proliferation assay

PBMCs resuspended in PBS were labeled with 0.5 γM CFSE (CellTrace CFSE cell proliferation kit, ThermoFisher) at room temperature for 8 minutes, followed by addition of RPMI 1640-10% FBS and extensive washing as described (Jenks et al., 2018; Kardava et al., 2018). The cells were then cultured with 2.5 γg/ml R848 and 1000 U/ml recombinant human IL-2 for 4 days at 1 × 10^6^ cells per well of a 48-well flat-bottom plate. The cells were collected and stained with mAbs against CD19, CD20, CD3, CD27, CD21, IgD, CD38, and fluorochrome-conjugated spike proteins RBD-B.1 and S1-B.1, fixed (Lysing Solution, BD Biosciences), permeabilized (Permeabilizing Solution 2; BD Biosciences) and stained with mAbs against IgG, IgA, IgM. The cells were acquired on an Aurora cytometer and the effect of stimulation on cell division was evaluated by analysis using FlowJo v10.

### Phosphorylation assay

PBMCs stained with mAbs against CD19, CD20, CD3, CD27, CD21, IgD, and BV421-conjugated RBD-B.1 were resuspended in RPMI 1640-10% FBS and stimulated with anti-BCR as described previously (Kardava et al., 2018), with the following modifications. The cells were stimulated at 37°C for 2 min with 10 µg/ml goat F(ab’)_2_ anti-human IgA/G/M (# 109-006-064 from Jackson ImmunoResearch Laboratories). For the detection of phosphorylated signaling intermediates, cells were fixed and permeabilized using BD Cytofix and Phosflow Perm/Wash buffers (BD Biosciences) and stained with Alexa Fluor 488-conjugated mAb against phosphorylated Syk (p-Y348) and Alexa Fluor 647-conjugated mAb against phosphorylated PLC-2 (p-Y759) (BD Biosciences). The samples were acquired on an Aurora cytometer and analysis was performed by FlowJo V10.

### Statistical analyses

Comparisons between groups and between timepoints within groups were performed by two-tailed non-parametric Mann-Whitney or Wilcoxon signed rank test respectively using Prism 9.4 (GraphPad) software. Comparisons between three timepoints were first performed by the Friedman test and if significant, pairwise Wilcoxon signed rank testing was then performed. Comparisons between the three groups were first performed by the Kruskal-Wallis test and if significant, pairwise Mann-Whitney testing was then performed. Associations between continuous variables were evaluated by Spearman rank order correlation. No adjustments for multiple comparisons were made for these analyses, thus the *P* values should be interpreted as suggestive rather than conclusive. Only significant *P* values (<0.05) are shown. Unless otherwise stated, the N for all data presented were 44 in the uninfected group and 11 each in the prior-infected and post-infected groups.

A secondary analysis used a linear mixed model regression applied to the base 10 logarithm of each outcome. The fixed effects were group (uninfected, prior-infected, and post-infected), days, and interactions between groups and days. Person-specific random deviations were allowed by the inclusion random intercepts. Because this parametric analysis tends to be less conservative than the nonparametric analyses described above, a sequentially rejective Bonferroni adjustment was made for the number of variants.

## Data Availability

All data produced in the present study are available upon reasonable request to the authors

## ACKNOWLEDGMENTS

We thank all participants for their willingness to take part in this study; Cathy Rehm, Ulisses Santamaria, Jessica Earhart, Kathleen Gittens and Michael Sneller for clinical support; Kaitlyn Sadtler and Dominic Esposito for development of antibody assays used in this study; Robin Carroll, Nazaire Jean-Baptiste and Lu Wang for technical assistance; and Robert Seder for helpful discussions. This work was funded by the Intramural Research Programs of the Vaccine Research Center and the Division of Intramural Research, National Institutes of Allergy and Infectious Diseases, National Institutes of Health.

## AUTHOR CONTRIBUTIONS

C.M.B., L.K., H.K., M.A.P., A.B.M, A.S.F., T.W.C., and S.M. designed the research. C.M.B., L.K., O.E.M., S.R.N, L.S., B.C.L, W.W., X.Z., F.L.A., S.E.M.K., G.E.M., L.H.P., H.K., M.A.P., A.B.M, T.W.C., and S.M. performed, analyzed and/or supervised the experiments. G.E.M., L.H.P., C.A.S., and M.A.R. oversaw clinical recruitment and/or medical aspects or assisted with sample collection. I.T.T. and P.D.K. provided critical reagents. L.K., T.W.C., and S.M. designed the figures. C.M.B, L.K., M.A.P., A.S.F., T.W.C., and S.M. wrote the manuscript. All authors reviewed and edited the manuscript.

## DECLARATION OF INTERESTS

The authors declare no competing interests.

## Notes

### Competing Interest Statement

The authors have declared no competing interest.

### Funding Statement

This study was funded by the Intramural Research Programs of the Vaccine Research Center and the Division of Intramural Research, National Institutes of Allergy and Infectious Diseases, National Institutes of Health.

### Author Declarations

The IRB of the National Institutes of Health gave ethical approval for this work

