## Supplemental Tables and Figures for "Recent SARS-CoV-2 infection abrogates antibody and B-cell responses to booster vaccination"

### SUPPLEMENTAL FIGURES AND TABLES

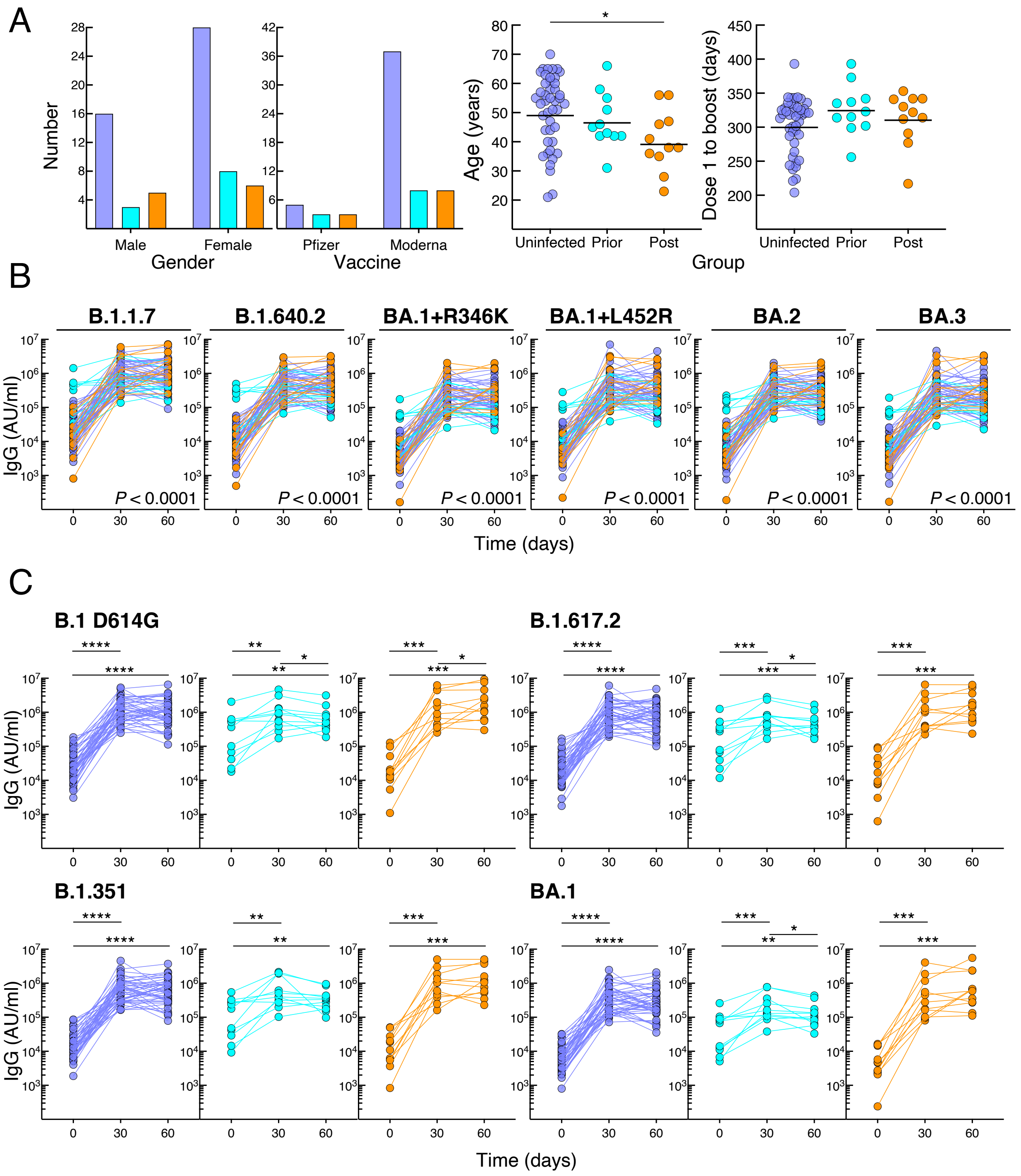

D

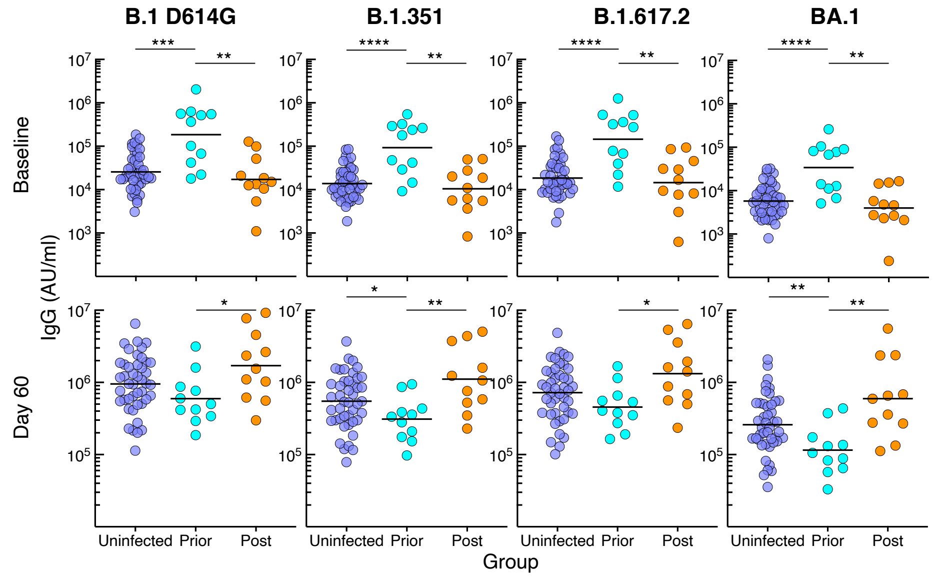
B.1 D614G
B.1.351
B.1.617.2
BA.1
Baseline
Day 60
IgG (AU/ml)
Uninfected
Prior
Post
Group

E

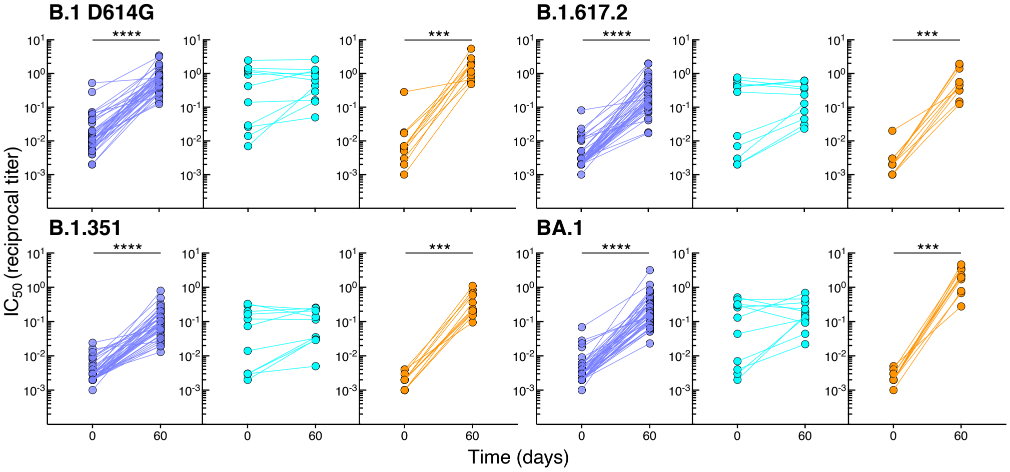
B.1 D614G
B.1.617.2
B.1.351
BA.1
IC50 (reciprocal titer)
Time (days)

F

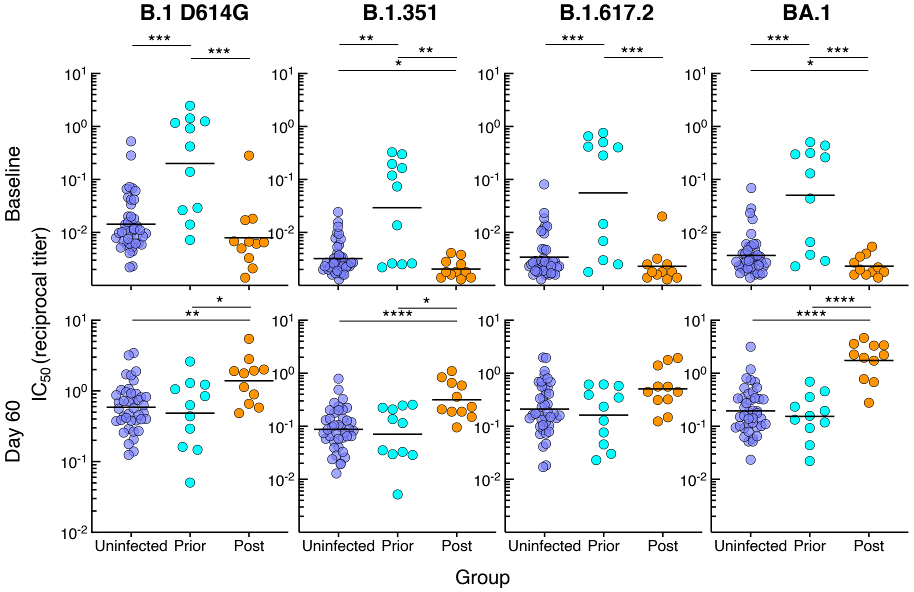
B.1 D614G
B.1.351
B.1.617.2
BA.1
Baseline
Day 60
IC50 (reciprocal titer)
Uninfected
Prior
Post
Group

**Figure S1. Demographics and antibody responses to booster vaccination**

(A) Comparison between the groups for gender, vaccine, age, and interval between dose 2 and booster vaccination.

(B) Serum IgG spike-binding titers for additional variants, expressed as in Figure 1B.

(C) Serum IgG spike-binding titers in Figure 1B shown by group.

(D) Comparison between the groups at baseline and day 60 of serum IgG spike-binding titers in Figure 1B.

(E) Serum neutralizing titers in Figure 1D shown by group.

(F) Comparison between the groups at baseline and day 60 of serum neutralizing titers in Figure 1D.

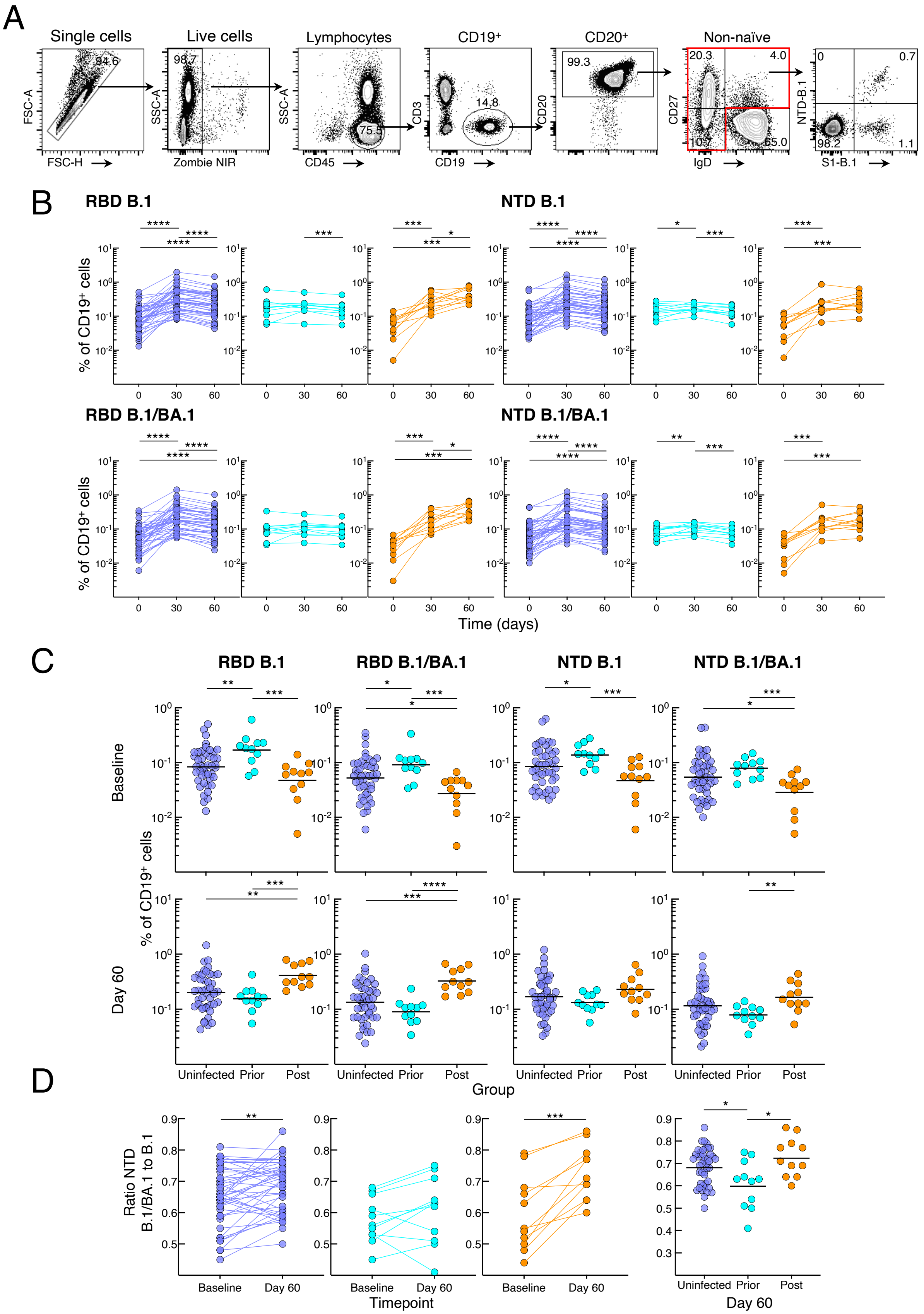
A
Single cells
Live cells
Lymphocytes
CD19⁺
CD20⁺
Non-naïve
FSC-A
FSC-H
SSC-A
Zombie NIR
CD45
CD3
CD19
CD20
CD27
IgD
NTD-B.1
S1-B.1
B
RBD B.1
NTD B.1
RBD B.1/BA.1
NTD B.1/BA.1
% of CD19⁺ cells
Time (days)
C
RBD B.1
RBD B.1/BA.1
NTD B.1
NTD B.1/BA.1
Baseline
Day 60
% of CD19⁺ cells
Uninfected Prior Post
Group
D
Ratio NTD B.1/BA.1 to B.1
Baseline Day 60
Timepoint
Uninfected Prior Post
Day 60

**Figure S2. Spike-binding B cells**

(A) Gating strategy for identifying non-naïve B cells; also showing representative binding of B.1 NTD and S1 tetramers.

(B) Frequencies of B.1 and BA.1 RBD and NTD tetramer-binding B cells in Figure 2B shown by group.

(C) Comparison between the groups at baseline and day 60 of frequencies of B.1 and BA.1 RBD and NTD tetramer-binding B cells in Figure 2B.

(D) As in Figure 2D but for NTD.

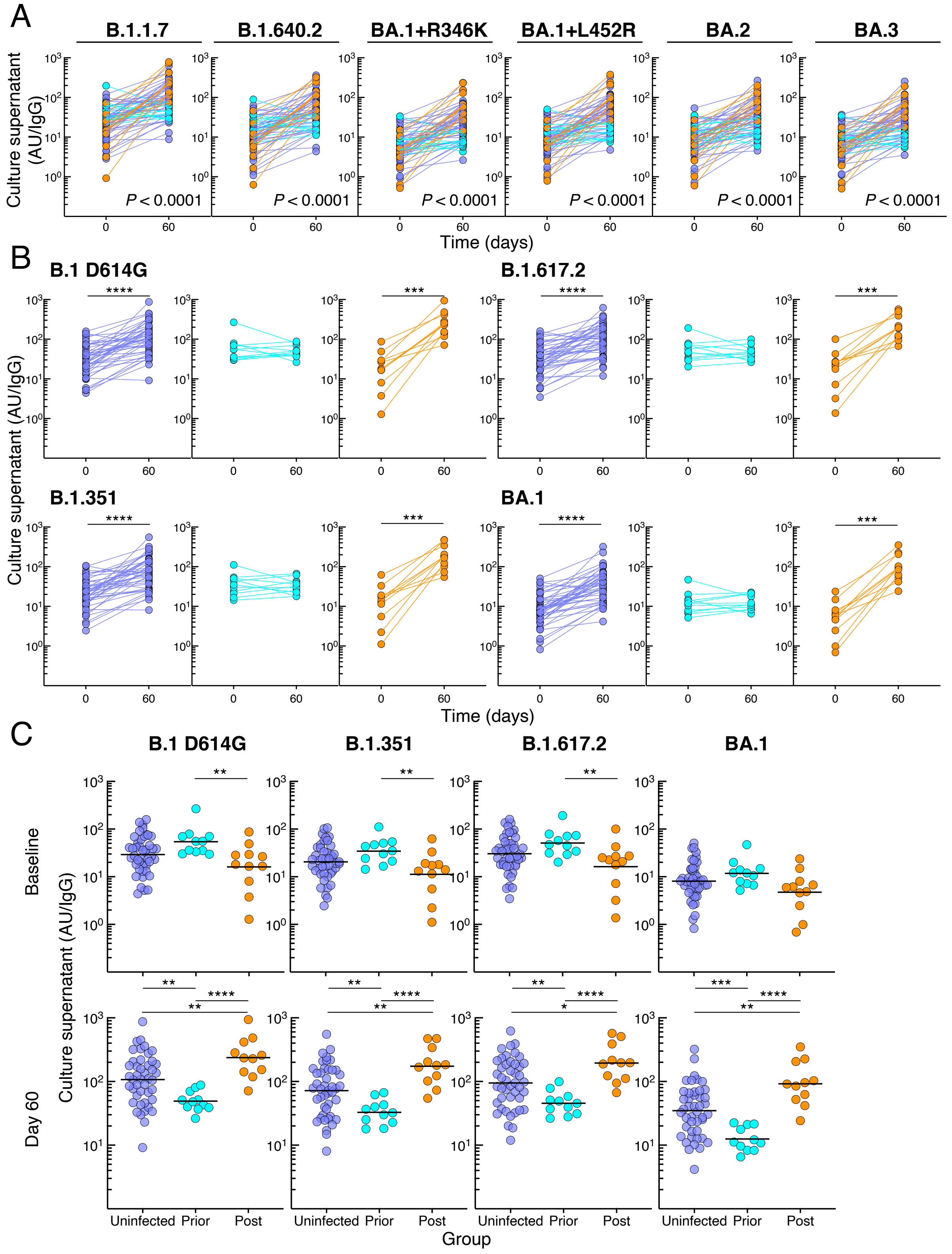
A
B.1.1.7
B.1.640.2
BA.1+R346K
BA.1+L452R
BA.2
BA.3
Culture supernatant (AU/IgG)
P < 0.0001
Time (days)
B
B.1 D614G
B.1.617.2
B.1.351
BA.1
C
Baseline
Day 60
Uninfected
Prior
Post
Group

**Figure S3. Spike-binding antibodies secreted from B cells in cultured PBMCs**

(A) Secreted IgG spike-binding titers for additional variants, expressed as in Figure 3A.

(B) Secreted IgG spike-binding titers in Figure 3A shown by group.

(C) Comparison between the groups at baseline and day 60 of secreted IgG spike-binding titers in Figure 3A.

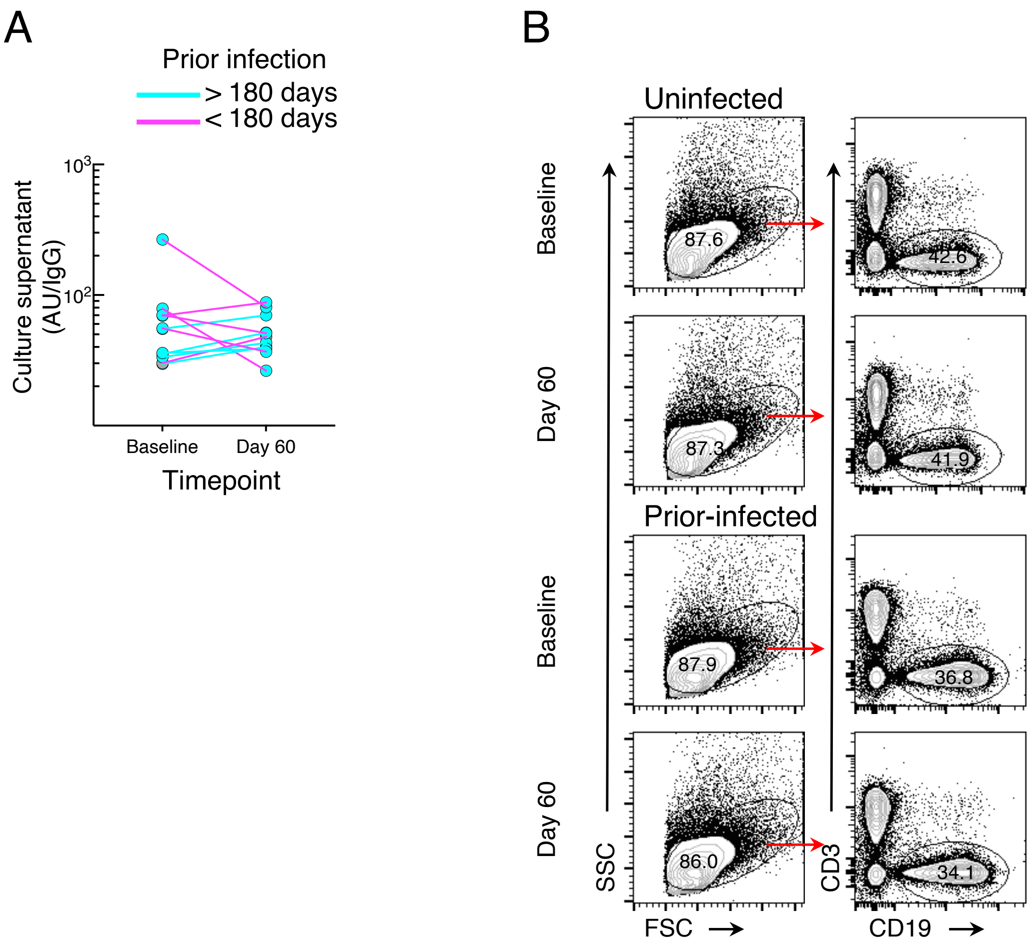


**Figure S4.** ***In vitro*** **proliferation studies**

(A) Secreted antibodies in Figure S3B for the prior-infected group against B.1 D614G spike shown color-coordinated by time of infection relative to booster vaccination. PBMCs of the six individuals with interval < 180 days were stimulated to evaluate cell division by CFSE dilution.

(B) Gating of live $CD19^{+}$ cells after four days in culture.

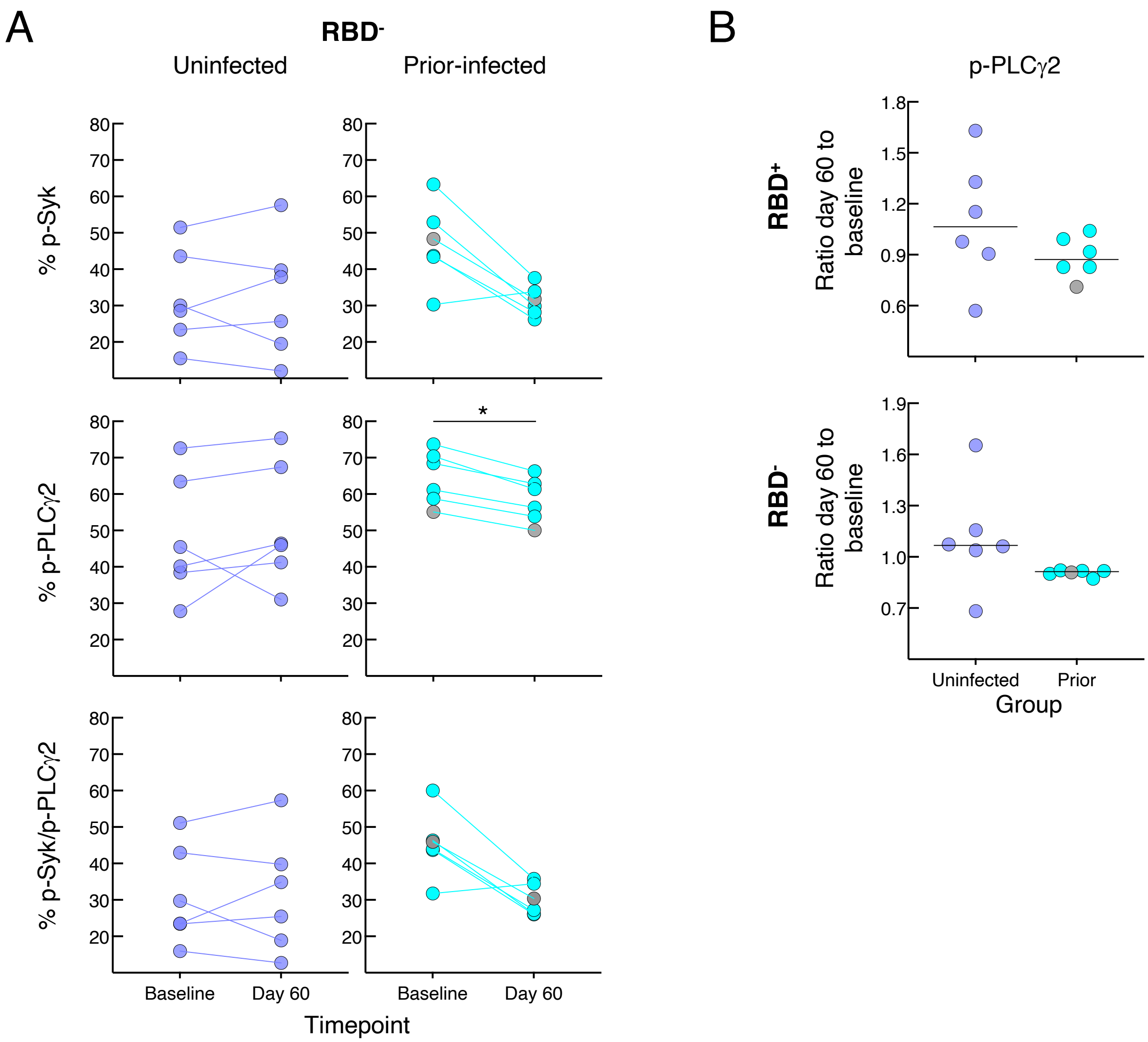


**Figure S5. *In vitro* BCR signaling studies**

(A) As in Figure 5B but for RBD⁻ B cells.

(B) As in Figure 5C but for single PLCγ2.

#### Table S1: *P* values for serum binding antibody increase over time or between group pairs

| Variant | Unin[a] day 0-30 | Unin day 0-60 | Unin day 30-60 | Prior infected day 0-30 | Prior infected day 0-60 | [b]Prior infected day 30-60 | Post infected day 0-30 | Post infected day 0-60 | Post infected day 30-60 | Prior vs unin day 0 | [c]Prior vs unin day 60 | Prior vs Post day 0 | [c]Prior vs Post day 60 | Post vs unin day 0 | Post vs unin day 60 | Day 60:0 fold unin vs prior | Day 60:0 fold post vs unin | Day 60:0 fold post vs prior |
|---|---|---|---|---|---|---|---|---|---|---|---|---|---|---|---|---|---|---|
| B.1.1.7 | <0.0001 | <0.0001 | ns | 0.002 | 0.002 | ns | 0.001 | 0.001 | 0.0186 | <0.0001 | ns | 0.0014 | 0.0128 | ns | ns | <0.0001 | 0.0080 | <0.0001 |
| B.1.640.2 | <0.0001 | <0.0001 | ns | 0.0029 | 0.0186 | 0.042 | 0.001 | 0.001 | 0.0322 | 0.0001 | ns | 0.0014 | 0.0052 | ns | 0.0457 | <0.0001 | 0.0060 | <0.0001 |
| BA.1 R346K | <0.0001 | <0.0001 | ns | 0.001 | 0.0049 | 0.0098 | 0.001 | 0.001 | ns | 0.0001 | 0.0065 | 0.0032 | 0.0024 | ns | ns | <0.0001 | 0.0174 | <0.0001 |
| BA.1 L452R | <0.0001 | <0.0001 | ns | 0.001 | 0.0029 | ns | 0.001 | 0.001 | ns | <0.0001 | 0.0164 | 0.0032 | 0.0024 | ns | ns | <0.0001 | 0.0222 | <0.0001 |
| BA.2 | <0.0001 | <0.0001 | ns | 0.002 | 0.0049 | 0.0322 | 0.001 | 0.001 | 0.0244 | <0.0001 | ns | 0.0014 | 0.0041 | ns | 0.0249 | <0.0001 | 0.0145 | <0.0001 |
| BA.3 | <0.0001 | <0.0001 | ns | 0.001 | 0.0049 | 0.0137 | 0.001 | 0.001 | ns | <0.0001 | 0.008 | 0.0024 | 0.0024 | ns | | <0.0001 | 0.0174 | <0.0001 |

[a]Uninfected

[b]Decreased from previous timepoint

[c]Decreased vs uninfected or post-infected

**Table S2: *P* values for group by time interaction for mixed-effects model of spike-binding serum antibody titers**

| Variant | All | Prior- vs post-infected | Uninfected vs prior-infected | Uninfected vs post-infected |
|---|---|---|---|---|
| B.1 D614G | <0.0001 | <0.0001 | <0.0001 | 0.1589 |
| B.1.1.7 | <0.0001 | <0.0001 | <0.0001 | 0.0706 |
| B.1.315 | <0.0001 | <0.0001 | <0.0001 | 0.1589 |
| B.1.617.2 | <0.0001 | <0.0001 | <0.0001 | 0.1589 |
| B.1.640.2 | <0.0001 | <0.0001 | <0.0001 | 0.0683 |
| BA.1 | <0.0001 | <0.0001 | <0.0001 | 0.0683 |
| BA.1 R346K | <0.0001 | <0.0001 | <0.0001 | 0.1034 |
| BA.1 L452R | <0.0001 | <0.0001 | <0.0001 | 0.1589 |
| BA.2 | <0.0001 | <0.0001 | <0.0001 | 0.1034 |
| BA.3 | <0.0001 | <0.0001 | <0.0001 | 0.1589 |

**Table S3: *P* Values for group by time interaction for mixed-effects model of frequencies of spike-binding B cells**

| Spike tetramer | All | Prior- vs post-infected | Uninfected vs prior-infected | Uninfected vs post-infected |
|---|---|---|---|---|
| B.1 S1 | <0.0001 | <0.0001 | <0.0001 | <0.0001 |
| B.1 RBD | <0.0001 | <0.0001 | <0.0001 | <0.0001 |
| B.1 NTD | <0.0001 | <0.0001 | <0.0001 | 0.0002 |
| B.1.& BA.1 RBD | <0.0001 | <0.0001 | <0.0001 | <0.0001 |
| B.1.& BA.1 NTD | <0.0001 | <0.0001 | <0.0001 | 0.0001 |

#### Table S4: *P* values for secreted antibody increase over time or between group pairs

| Variant | Unin[a] day 0-36 | Prior infected day 0-60 | Post infected day 0-60 | Prior vs unin day 0 | [b]Prior vs unin day 60 | Prior vs post day 0 | [b]Prior vs post day 60 | Post vs unin day 0 | Post vs unin day 60 | Day 60:0 fold unin vs prior | Day 60:0 fold post vs unin | Day 60:0 fold post vs prior |
|---|---|---|---|---|---|---|---|---|---|---|---|---|
| B.1.1.7 | <0.0001 | ns | 0.001 | 0.0154 | 0.007 | 0.0024 | <0.0001 | ns | 0.0029 | <0.0001 | 0.0002 | <0.0001 |
| B.1.640.2 | <0.0001 | ns | 0.001 | 0.0220 | 0.006 | 0.0052 | <0.0001 | ns | 0.0045 | <0.0001 | 0.0004 | <0.0001 |
| BA.1 R346K | <0.0001 | ns | 0.001 | ns | 0.0001 | ns | <0.0001 | ns | 0.0060 | <0.0001 | 0.0001 | <0.0001 |
| BA.1 L452R | <0.0001 | ns | 0.001 | ns | 0.0006 | ns | <0.0001 | ns | 0.0039 | <0.0001 | 0.0002 | <0.0001 |
| BA.2 | <0.0001 | ns | 0.001 | ns | 0.0031 | ns | <0.0001 | ns | 0.0031 | <0.0001 | 0.0007 | <0.0001 |
| BA.3 | <0.0001 | ns | 0.001 | ns | 0.0006 | ns | <0.0001 | ns | 0.0033 | <0.0001 | 0.0001 | <0.0001 |

[a]Uninfected

[b]Decreased vs uninfected or post-infected

**Table S5: *P* values for group by time interaction for mixed-effects model of spike-binding secreted antibody titers**

| Variant | All | Prior- vs post-infected | Uninfected vs prior-infected | Uninfected vs post-infected |
|---|---|---|---|---|
| B.1 D614G | <0.0001 | <0.0001 | <0.0001 | 0.0001 |
| B.1.1.7 | <0.0001 | <0.0001 | <0.0001 | <0.0001 |
| B.1.315 | <0.0001 | <0.0001 | <0.0001 | 0.0001 |
| B.1.617.2 | <0.0001 | <0.0001 | <0.0001 | 0.0001 |
| B.1.640.2 | <0.0001 | <0.0001 | <0.0001 | 0.0001 |
| BA.1 | <0.0001 | <0.0001 | <0.0001 | <0.0001 |
| BA.1 R346K | <0.0001 | <0.0001 | <0.0001 | 0.0001 |
| BA.1 L452R | <0.0001 | <0.0001 | <0.0001 | 0.0001 |
| BA.2 | <0.0001 | <0.0001 | <0.0001 | 0.0001 |
| BA.3 | <0.0001 | <0.0001 | <0.0001 | 0.0001 |

### Table S6: 21-color flow cytometry panel

| Reagent | Source |
|---|---|
| Mouse anti-human CD45 BUV805 (clone HI30) | BD Biosciences |
| Mouse anti-human CD19 BV650 (Clone SJ25-C1) | BD Biosciences |
| Mouse anti-human CD20 APC-H7 (Clone 2H7) | BD Biosciences |
| Mouse anti-human CD10 BV510 (Clone HI10a) | BD Biosciences |
| Mouse anti-human IgG PE-Cy7 (Clone G18-145) | BD Biosciences |
| Mouse anti-human CD11c BUV395 (clone B-Ly6) | BD Biosciences |
| Mouse anti-human CD3 BV570 (Clone UCHT1) | Biolegend |
| Mouse anti-human IgD BV605 (Clone IA6-2) | Biolegend |
| Mouse anti-human IgM BV711 (Clone MHM-88) | Biolegend |
| Mouse anti-human CD27 BV785 (Clone O323) | Biolegend |
| Mouse anti-human CD21 PE/Dazzle594 (Clone BU32) | Biolegend |
| Mouse anti-human CD38 APC/Fire810 (Clone HB-7) | Biolegend |
| Mouse anti-human CD71 Alexa Fluor 700 (Clone CY1G4) | Biolegend |
| Mouse anti-human IgA VioBlue (Clone IS11-8E10) | Miltenyi Biotec |
| SARS-CoV-2 S1 B.1 (#793806) | Biolegend |
| SARS-CoV-2 Spike B.1 trimer (#SPN-C82E9) | AcroBiosystems |
| SARS-CoV-2 RBD B.1 (#SPD-C82E9) | AcroBiosystems |
| SARS-CoV-2 RBD BA.1 (#SPD-C82E4) | AcroBiosystems |
| SARS-CoV-2 S NTD B.1 | In-house |
| SARS-CoV-2 S NTD BA.1 | In-house |
| Streptavidin PE | ThemoFisher |
| Streptavidin APC | ThemoFisher |
| Streptavidin PE-Cy5.5 | ThemoFisher |
| Streptavidin Alexa Fluor 488 | ThemoFisher |
| Streptavidin BV421 | Biolegend |
| Streptavidin BUV615 | BD Biosciences |
| Zombie NIR Fixable Viability Dye | Biolegend |
